# Epidemiology and Survival of Primary Extraosseous Plasmacytoma: Insights from A Population-Based Study with A 20-Year Follow-Up

**DOI:** 10.1101/2023.05.02.23289425

**Authors:** Fan Wang

**Affiliations:** Department of Hematology, Tongji Hospital, Tongji Medical College, Huazhong University of Science and Technology, Wuhan, Hubei, The People’s Republic of China

**Keywords:** extraosseous plasmacytoma, SEER, survival, outcome, nomogram.

## Abstract

**Background:** Primary extraosseous plasmacytoma (PEP) is a rare and localized form of plasmacytoma that is not well understood. This study aimed to investigate the clinical features and prognostic factors associated with PEP.

**Methods:** Using the Surveillance, Epidemiology, and End Results (SEER) database, patients diagnosed with PEP between 2000 and 2019 were extracted. Survival curves were calculated with the Kaplan-Meier method, and prognostic factors were identified based on the univariate and multivariate Cox regression analyses. A nomogram was constructed to predict survival at 3 and 5 years after diagnosis.

**Results:** A total of 1044 PEP patients were included in this study. The average age was 60.3 ± 15.2 years, with 64.3% being male (male: female = 1.8:1) and 53.8% being over 60 years old. The most affected sites were the upper aerodigestive tract (42.6%). Most patients received radiotherapy (60.7%), followed by surgery (52.8%) and chemotherapy (15.1%). Overall survival (OS) rates were 88.3% at 1 year, 78.1% at 3 years, 70.7% at 5 years, and 56.0% at 10 years. Corresponding cancer-specific survival (CSS) rates were 92.1%, 85.8%, 82.5%, and 78.1%, respectively. Multivariate cox analysis indicated that age, race, marital status at diagnosis, radiotherapy, chemotherapy, and surgery were independent prognostic factors for OS, while age, race, radiotherapy, chemotherapy, and surgery were independent prognostic factors for CSS. Nomograms were further constructed to predict the possibility of OS and CSS with good performances.

**Conclusions:** The survival outcome of patients with PEP depends on several factors including age, race, marital status, and treatment options such as chemotherapy, radiotherapy, and surgery, which were also identified as independent predictors of OS for PEP. Patients who were younger, Asian or Pacific Islander, American Indian or Native American, and received radiotherapy or surgery had a more favorable prognosis, while those who underwent chemotherapy had poorer outcomes. This study provides valuable insights into the management of PEP.

## Introduction

Extraosseous plasmacytoma (EOP) is a rare type of plasma cell neoplasm that occurs outside the bone marrow^1, 2^. It is characterized by the accumulation of malignant plasma cells in the soft tissues of the body, such as the skin, lymph nodes, and organs including the lung, bladder, respiratory tract, and gastrointestinal tract^3^. EOP accounts for less than 5% of all plasma cell neoplasms, with over 80% occurring in the upper aerodigestive tract (UAD) ^4^. It typically affects older individuals, with a higher incidence in men than women (male-to-female ratio of 1.7:1) ^5^. PEP usually has an indolent clinical course, responds well to local radiotherapy, but has a tendency to local relapse, and rarely progresses to multiple myeloma (MM) ^6^.

Diagnosis and management of EOP still pose a challenge due to its rarity and lack of specific symptoms^7^. A detailed medical history and physical examination are usually the initial steps, ^8^ followed by imaging tests like X-rays, CT scans, MRI scans, or PET scans to identify the tumor’s presence and location^9^. To confirm the diagnosis, a biopsy of the suspected tumor is necessary to be performed, and in some cases, a bone marrow biopsy and aspiration may also be conducted to exclude multiple myeloma^1^. Clonal plasma cells’ demonstration in the biopsy sample confirms the diagnosis, and immunohistochemical staining and flow cytometry may also be performed to further characterize the neoplasm and distinguish it from other plasma cell neoplasms^7, 10^.

The prognosis for EOP depends on the location and extent of the disease, with patients who have isolated EOP having a better outcome than those with disseminated disease^11^. Therapy options include surgery, radiation therapy, and systemic therapy such as chemotherapy^6^. The goal of therapy is to remove abnormal plasma cells and prevent metastasis^6^. Surgery is the most common therapy option, but radiation therapy may also be used as the primary treatment or in combination with surgery or chemotherapy^4, 12^. The outcome varies depending on several factors, with early-stage EOP patients having a better prognosis than those with advanced stages of the disease^13^. The 5-year survival rate is approximately 90% for early-stage EOP patients who undergo complete surgical removal of the tumor^14^. However, patients with more advanced stages or recurrent EOP may require more intensive treatment, including combination therapy with radiation therapy, chemotherapy, or stem cell transplantation^15^. In some cases, EOP may progress to multiple myeloma, a more advanced and potentially life-threatening form of plasma cell cancer^16^.

However, EOP, especially primary extraosseous plasmacytoma (PEP) is not well understood owing to its rarity. The purpose of this study is to identify the factors affecting the survival of PEP patients based on a population-based study using the national Cancer Institute’s Surveillance, Epidemiology and End Results (SEER) database.

## Materials and Methods

### Data acquisition

Data used for the current study were acquired from the Surveillance, Epidemiology, and End Results (SEER) Program (https://seer.cancer.gov/), which is maintained by the National Cancer Institute (NCI). The data were collected using SEER*Stat software version 8.4.0.1 (https://seer.cancer.gov/seerstat/, accessed on November 25, 2022). Patients with extraosseous plasmacytoma who were diagnosed between 2000 and 2019 were selected from the “Incidence-SEER Research Plus Data, 17 Registries, Nov 2021 Sub (2000-2019)” using the case listing session. Only cases with known age (censored at age 89 years) and only malignant behavior were included. The inclusion criteria were as follows: (1) the International Classification of Diseases for Oncology (ICD-O-3) histologic code (9734/3); (2) the sequence number indicating the only primary or first primary. The exclusion criteria were as follows: (1) the type of reporting source was “death certificate only” or “autopsy only”; (2) the patient’s survival time was 0 or unknown; (3) patients with bone marrow or “unknown” site as the involvement of primary site; (4) the diagnosis confirmation was “unknown” and the marital status was “unknown”. A total of 1044 patients with primary extraosseous plasmacytoma were included in the final cohort. This study used publicly available SEER data, and because the patients’ private information was anonymous and cannot be reidentified, ethical approval from an ethics committee was not required. The flow chart in Figure 1 shows the selection process.

**Figure 1.**
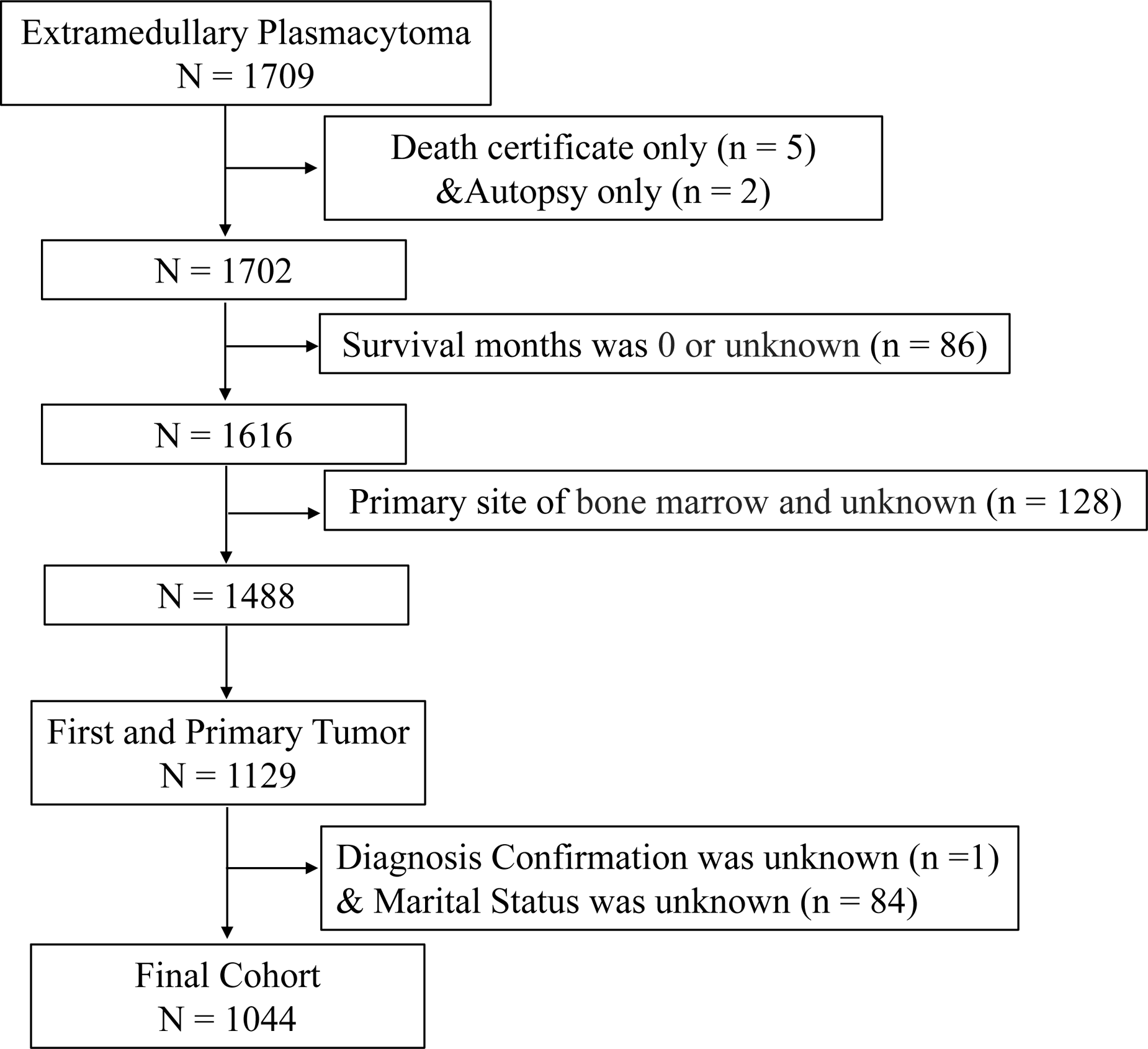
Flow chart of study cohort selection using the SEER database. A flow diagram of primary extraosseous plasmacytoma patient selection in this study. SEER, Surveillance, Epidemiology, and End Results.

### Definition of Variables

The analysis involved including a range of variables such as age, sex, race, marital status, year of diagnosis, primary site, vital status, survival months, COD to site recode, cause-specific death classification, cause of death to site, sequence number, first malignant primary indicator, total number of in situ/malignant tumors for patient, type of reporting source, diagnostic confirmation, surgery of primary site, chemotherapy recode, and radiation recode. Age was classified into two groups, namely < 60 years old and 60 + years old, with reference to the age at diagnosis of extraosseous plasmacytoma. Race was divided into African American, White, and Other (comprising “Asian/Pacific Islander” and “American Indian/Alaska Native”).

Marital status was grouped as married, single, and other (including “Divorced”, “Separated”, and “Widowed”). Cause-of-death information was obtained from the “COD to site recode” field. Extraosseous plasmacytoma-related death was defined in the SEER database as “dead (attributed to this cancer diagnosis)”. Diagnosis years were categorized as “2000-2009” and “2010-2019”. The definition of extraosseous plasmacytoma-unrelated death was defined in the SEER database as death “dead (attributable to causes other than this cancer diagnosis)”. The overall survival time was calculated from the date of extraosseous plasmacytoma diagnosis to death or last follow up.

### Statistical analysis

The R program language (http://www.r-project.org/, version 4.2.1; R Foundation for Statistical Computing, Vienna, Austria) was used for all statistical analyses in this study. Two groups, “extraosseous plasmacytoma-related death” and “other cause,” were compared in terms of baseline characteristics of primary extraosseous plasmacytoma patients. Student’s t-test and chi-square test were used to compare continuous and categorical data, respectively. Kaplan-Meier method was used to estimate overall survival (OS) and cancer-specific survival (CSS) with log-rank tests to evaluate the differences between groups. Univariate and multivariate Cox proportional regression analyses were conducted to identify independent prognostic factors of primary extraosseous plasmacytoma patients. Hazard ratios (HR) estimates and 95% confidence intervals (95% CIs) were reported. The Cox proportional hazards regression model was evaluated for proportionality assumptions using preliminary tests, which revealed no violations of any variables. Variables with *P*LJ<LJ0.10 in the univariate Cox proportional hazard model were included in the multivariate Cox model. Variables with *P*LJ<LJ0.05 in the multivariate model were considered significant. Based on independent prognostic factors from the multivariate Cox analysis, nomograms were constructed to predict the 3- and 5-year OS and CSS probabilities. Time-dependent receiver operating characteristic (ROC) curves of nomograms were generated for 3- and 5-year time points, and the corresponding area under the curve (AUC) was used to assess the discrimination. Calibration curves and decision curve analysis (DCA) nomograms were plotted to evaluate the performance of the nomogram at 3- and 5-year time points. All *P* values were two-sided, and a *P* value < 0.05 was considered statistically significant.

## Results

### Characteristics of Primary Extraosseous Plasmacytoma Patients at Baseline

A total of 1044 patients diagnosed with primary extraosseous plasmacytoma in the SEER 17 registry, Nov 2021 Sub (2000-2019) from January 2000 to December 2019 were analyzed, with their baseline characteristics presented in Figure 1. The upper aerodigestive tract was the most affected site, accounting for 42.6% (*n* = 445) of cases, while other sites such as the digestive system, nervous system, respiratory system, lymph nodes, and other areas were also affected. Table 1 provides a detailed description of the primary site distribution. Of the study cohort, 64.3% were male (male: female = 1.8:1), and the average age at diagnosis was 60.3 ± 15.2 years, with 46.2% under the age of 60 (< 60) and 53.8% aged 60 or older (60+). Most of the patients were white (79.8%), followed by African American and other racial groups (11.3% and 8.9%, respectively). At diagnosis, 64.8% of patients were married, 19.0% had other marital statuses (such as divorced, separated, or widowed), and 16.3% were single who had never been married. The highest incidence of primary extraosseous plasmacytoma occurred in the 2010-2019 period (57.2%). The majority of patients (60.7%) received radiotherapy, while 52.8% were treated with surgery and 15.1% received chemotherapy. At the time of the last follow-up, 62.1% of patients were still alive, while 18.2% had died from primary extraosseous plasmacytoma and an additional 19.7% had died from other causes, such as heart disease, cerebrovascular disease, and kidney disease. Notably, only chemotherapy showed a significant difference (*P* < 0.001) between the two survival groups: “extraosseous plasmacytoma-related death” and “other cause”. A summary of the epidemiologic characteristics and survival comparison were shown in Table 2.

**Table 1.** Distribution of the primary sites of extraosseous plasmacytoma

| Primary Sites | Overall<br>(N=1044) |
| --- | --- |
| Upper aerodigestive tract | 445 (42.6%) |
| Digestive system | 101 (9.7%) |
| Nervous system | 40 (3.8%) |
| Respiratory system | 70 (6.7%) |
| Skin | 31 (3.0%) |
| Urinary System | 30 (2.9%) |
| Reproductive System | 34 (3.3%) |
| Lymph nodes | 60 (5.7%) |
| Other | 233 (22.3%) |

**Table 2.**
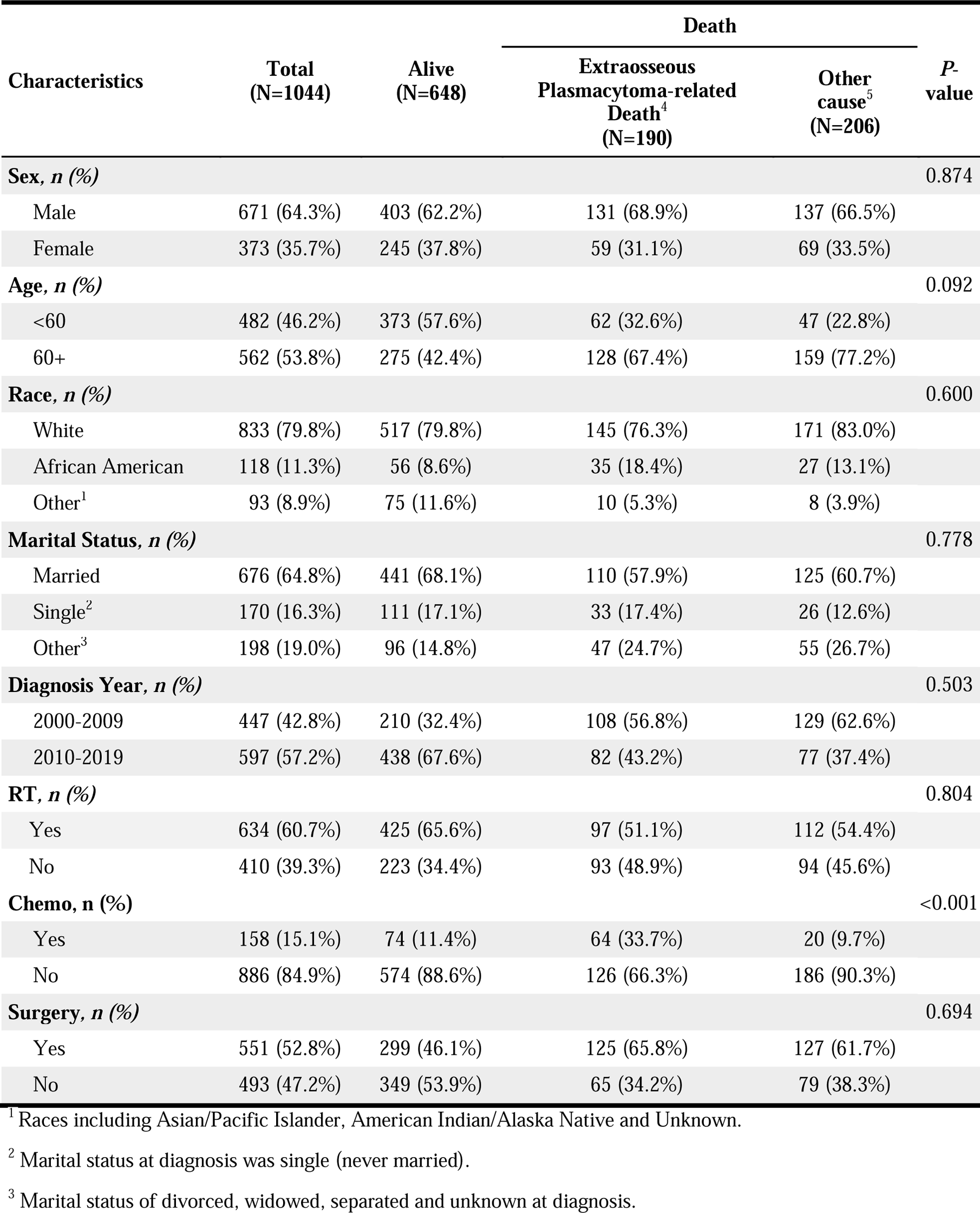
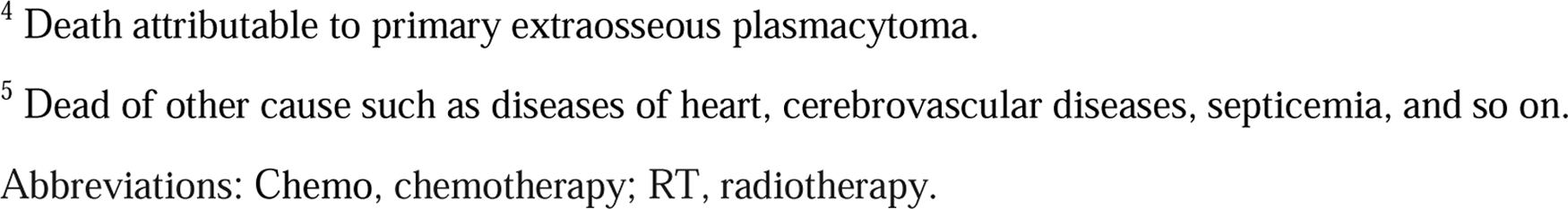
Comparison of patient baseline characteristics between extraosseous plasmacytoma-related death and other cause in the primary extraosseous plasmacytoma cohort.

| Characteristics | Total<br>(N=1044) | Alive<br>(N=648) | Death |  | P-value |
| --- | --- | --- | --- | --- | --- |
|  |  |  | Extraosseous<br>Plasmacytoma-related<br>Death<sup>4</sup><br>(N=190) | Other<br>cause<sup>5</sup><br>(N=206) |  |
| **Sex, n (%)** |  |  |  |  | 0.874 |
| Male | 671 (64.3%) | 403 (62.2%) | 131 (68.9%) | 137 (66.5%) |  |
| Female | 373 (35.7%) | 245 (37.8%) | 59 (31.1%) | 69 (33.5%) |  |
| **Age, n (%)** |  |  |  |  | 0.092 |
| <60 | 482 (46.2%) | 373 (57.6%) | 62 (32.6%) | 47 (22.8%) |  |
| 60+ | 562 (53.8%) | 275 (42.4%) | 128 (67.4%) | 159 (77.2%) |  |
| **Race, n (%)** |  |  |  |  | 0.600 |
| White | 833 (79.8%) | 517 (79.8%) | 145 (76.3%) | 171 (83.0%) |  |
| African American | 118 (11.3%) | 56 (8.6%) | 35 (18.4%) | 27 (13.1%) |  |
| Other<sup>1</sup> | 93 (8.9%) | 75 (11.6%) | 10 (5.3%) | 8 (3.9%) |  |
| **Marital Status, n (%)** |  |  |  |  | 0.778 |
| Married | 676 (64.8%) | 441 (68.1%) | 110 (57.9%) | 125 (60.7%) |  |
| Single<sup>2</sup> | 170 (16.3%) | 111 (17.1%) | 33 (17.4%) | 26 (12.6%) |  |
| Other<sup>3</sup> | 198 (19.0%) | 96 (14.8%) | 47 (24.7%) | 55 (26.7%) |  |
| **Diagnosis Year, n (%)** |  |  |  |  | 0.503 |
| 2000-2009 | 447 (42.8%) | 210 (32.4%) | 108 (56.8%) | 129 (62.6%) |  |
| 2010-2019 | 597 (57.2%) | 438 (67.6%) | 82 (43.2%) | 77 (37.4%) |  |
| **RT, n (%)** |  |  |  |  | 0.804 |
| Yes | 634 (60.7%) | 425 (65.6%) | 97 (51.1%) | 112 (54.4%) |  |
| No | 410 (39.3%) | 223 (34.4%) | 93 (48.9%) | 94 (45.6%) |  |
| **Chemo, n (%)** |  |  |  |  | <0.001 |
| Yes | 158 (15.1%) | 74 (11.4%) | 64 (33.7%) | 20 (9.7%) |  |
| No | 886 (84.9%) | 574 (88.6%) | 126 (66.3%) | 186 (90.3%) |  |
| **Surgery, n (%)** |  |  |  |  | 0.694 |
| Yes | 551 (52.8%) | 299 (46.1%) | 125 (65.8%) | 127 (61.7%) |  |
| No | 493 (47.2%) | 349 (53.9%) | 65 (34.2%) | 79 (38.3%) |  |

<sup>1</sup> Races including Asian/Pacific Islander, American Indian/Alaska Native and Unknown.
<sup>2</sup> Marital status at diagnosis was single (never married).
<sup>3</sup> Marital status of divorced, widowed, separated and unknown at diagnosis.
<sup>4</sup> Death attributable to primary extraosseous plasmacytoma.
<sup>5</sup> Dead of other cause such as diseases of heart, cerebrovascular diseases, septicemia, and so on.
Abbreviations: Chemo, chemotherapy; RT, radiotherapy.

### Analysis of Survival for Patients with Primary Extraosseous Plasmacytomaa

During the follow-up period, 396 deaths occurred, with 190 of those being specific to PEP. As shown in Figure 2A and B, the 1-year, 3-year, 5-year, and 10-year overall survival (OS) rates were 88.3%, 78.1%, 70.7%, and 56.0%, respectively, and the corresponding cancer-specific survival (CSS) rates were 92.1%, 85.8%, 82.5%, and 78.1%, respectively. There were no significant differences observed between the two diagnosis year groups “2000-2009” and “2010-2019” (*P* = 0.29; Figure 2C). Neither were there significant differences observed for CSS between the diagnosis years (*P* = 0.14; Figure 2D). Furthermore, Kaplan-Meier analysis of OS suggested that age over 60 years old (*P* < 0.0001; Figure 3A), being African American (Figure 3C), having a marital status of “other” (*P* < 0.0001 vs. “Married” and *P* = 0.001 vs. “Single”; Figure 3D), receiving chemotherapy (*P* < 0.0001; Figure 3E), not receiving radiotherapy (*P* < 0.0001; Figure 3F), and not undergoing surgery (*P* < 0.0001; Figure 3G) were all associated with poorer overall survival. There was no significant difference in overall survival between the sex categories (*P* = 0.11, Figure 3B). Moreover, Figure 4 showed that Kaplan-Meier analysis of CSS yielded similar results.

**Figure 2.**
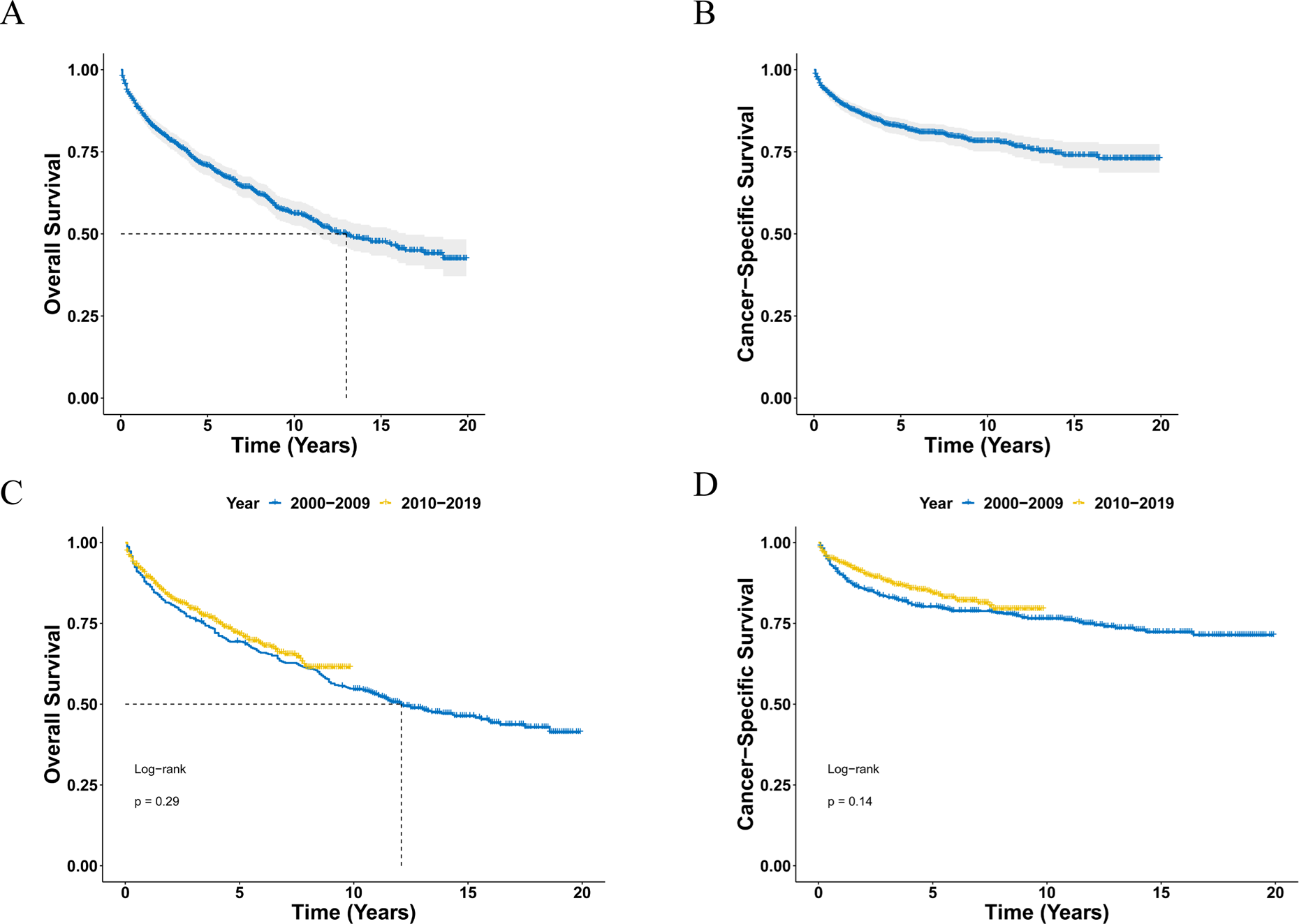
Survival analysis of primary extraosseous plasmacytoma. **(A, B)** OS (A) and CSS (B) curves for all primary extraosseous plasmacytoma patients. **(C, D)** Survival curves of OS (C) and CSS (D) according to the diagnosis years. OS, overall survival; CSS, cancer-specific survival.

**Figure 3.**
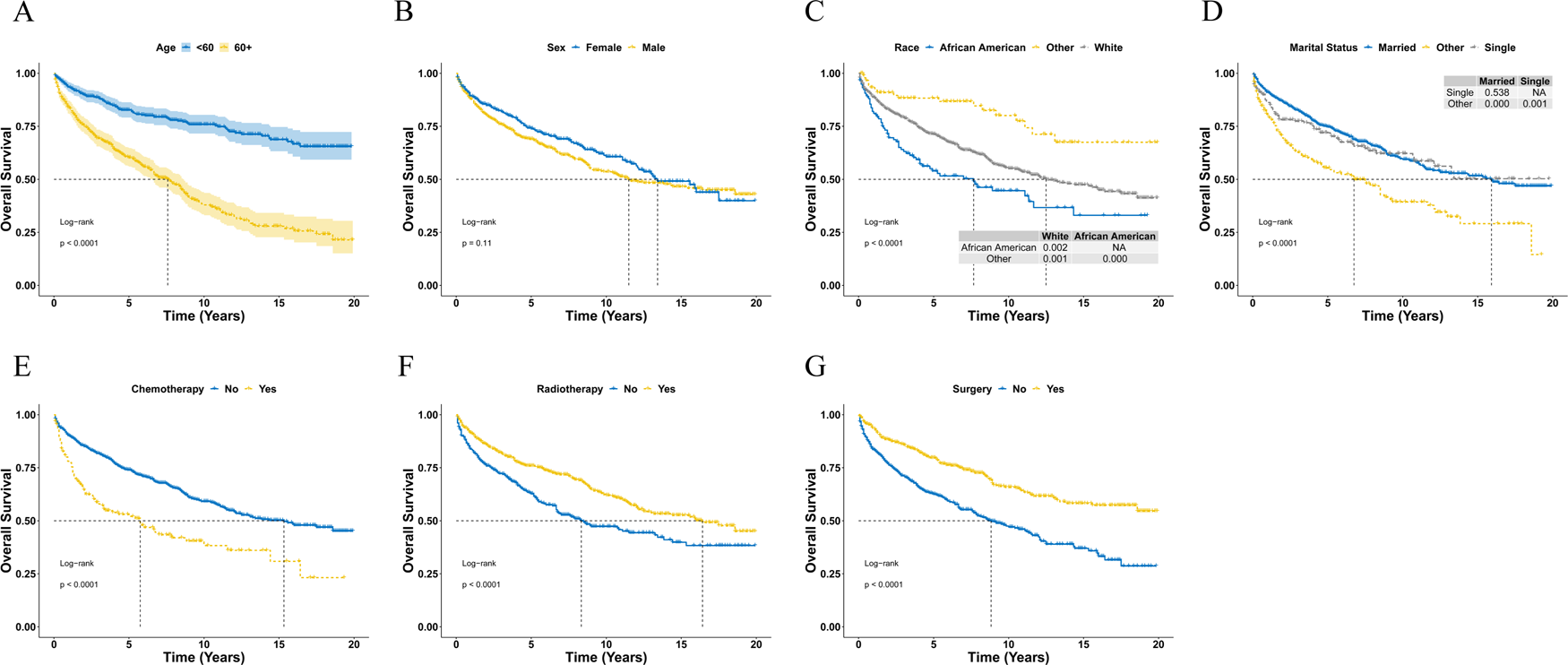
Overall survival analysis of primary extraosseous plasmacytoma stratified by age (A), sex (B), race (C), and marital status (D), chemotherapy (E), radiotherapy (F) and surgery (G) using Kaplan-Meier method. NA, not applicable.

**Figure 4.**
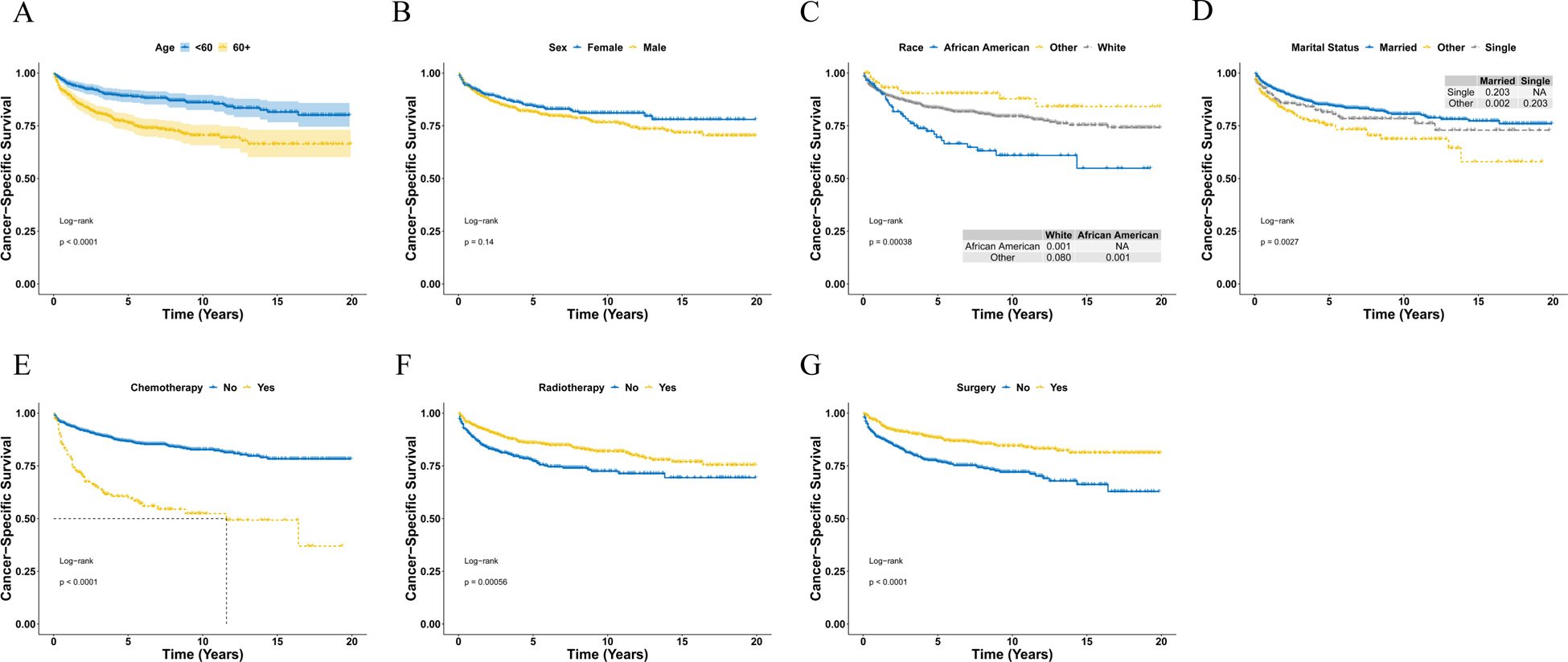
Cancer-specific survival analysis of primary extraosseous plasmacytoma stratified by age (A), sex (B), race (C), and marital status (D), chemotherapy (E), radiotherapy (F) and surgery (G) using Kaplan-Meier method. NA, not applicable.

### Univariate and Multivariable Cox Regression Analysis of Primary Extraosseous Plasmacytoma Patients

Univariate Cox regression analysis of overall survival (OS) revealed that age, race, marital status at diagnosis, radiotherapy, chemotherapy, and surgery were significantly associated with OS (Table 3). Conversely, variables such as sex and diagnosis year had no effect on the OS outcomes of primary extraosseous plasmacytoma. The univariate Cox regression analysis of cancer-specific survival (CSS) produced similar results (Table 3). The significant variables were subsequently included in the multivariate Cox analysis, which indicated that age, race, marital status at diagnosis, radiotherapy, chemotherapy, and surgery were independent prognostic factors for OS (Table 4). Concerning CSS, age, race, radiotherapy, chemotherapy, and surgery were identified as independent prognostic factors (Table 4).

**Table 3.** Univariable cox regression analysis for overall survival of primary extraosseous plasmacytoma patients

| Parameters | OS |  |  | CSS |  |  |
| --- | --- | --- | --- | --- | --- | --- |
|  | HR | 95% CI | P-value | HR | 95% CI | P-value |
| **Sex** |  |  |  |  |  |  |
| Female | *ref* |  |  | *ref* |  |  |
| Male | 1.19 | [0.96, 1.47] | 0.109 | 1.26 | [0.92, 1.71] | 0.144 |
| **Age** |  |  |  |  |  |  |
| <60 | *ref* |  |  | *ref* |  |  |
| 60+ | 3.12 | [2.50, 3.90] | < 0.001 | 2.22 | [1.64, 3.01] | < 0.001 |
| **Race** |  |  |  |  |  |  |
| White | *ref* |  |  | *ref* |  |  |
| African American | 1.54 | [1.17, 2.02] | 0.002 | 1.84 | [1.28, 2.67] | 0.001 |
| Other<sup>1</sup> | 0.45 | [0.28, 0.72] | < 0.001 | 0.57 | [0.30, 1.08] | 0.082 |
| **Marital Status** |  |  |  |  |  |  |
| Married | *ref* |  |  | *ref* |  |  |
| Single<sup>2</sup> | 1.10 | [0.82, 1.46] | 0.529 | 1.29 | [0.87, 1.90] | 0.201 |
| Other<sup>3</sup> | 1.93 | [1.53, 2.44] | < 0.001 | 1.80 | [1.28, 2.54] | < 0.001 |
| **Diagnosis Year** |  |  |  |  |  |  |
| 2000-2009 | *ref* |  |  | *ref* |  |  |
| 2010-2019 | 0.89 | [0.72, 1.10] | 0.290 | 0.80 | [0.59, 1.08] | 0.143 |
| **RT** |  |  |  |  |  |  |
| No | *ref* |  |  | *ref* |  |  |
| Yes | 0.63 | [0.51, 0.76] | < 0.001 | 0.61 | [0.46, 0.81] | < 0.001 |
| **Chemo** |  |  |  |  |  |  |
| No | *ref* |  |  | *ref* |  |  |
|  | HR | 95% CI | P-value | HR | 95% CI | P-value |
| Yes | 2.06 | [1.62, 2.62] | < 0.001 | 3.60 | [2.66, 4.88] | < 0.001 |
| **Surgery** |  |  |  |  |  |  |
| No | *ref* |  |  | *ref* |  |  |
| Yes | 0.51 | [0.41, 0.62] | < 0.001 | 0.49 | [0.36, 0.66] | < 0.001 |

<sup>1</sup> Races including Asian/Pacific Islander and American Indian/Alaska Native.
<sup>2</sup> Marital status at diagnosis was single (never married).
<sup>3</sup> Marital status of divorced, widowed and separated at diagnosis.
Abbreviations: Chemo, chemotherapy; CI, confidence interval; CSS, cancer-specific survival; HR, hazard ration; OS, overall survival; RT, radiotherapy.

**Table 4.**
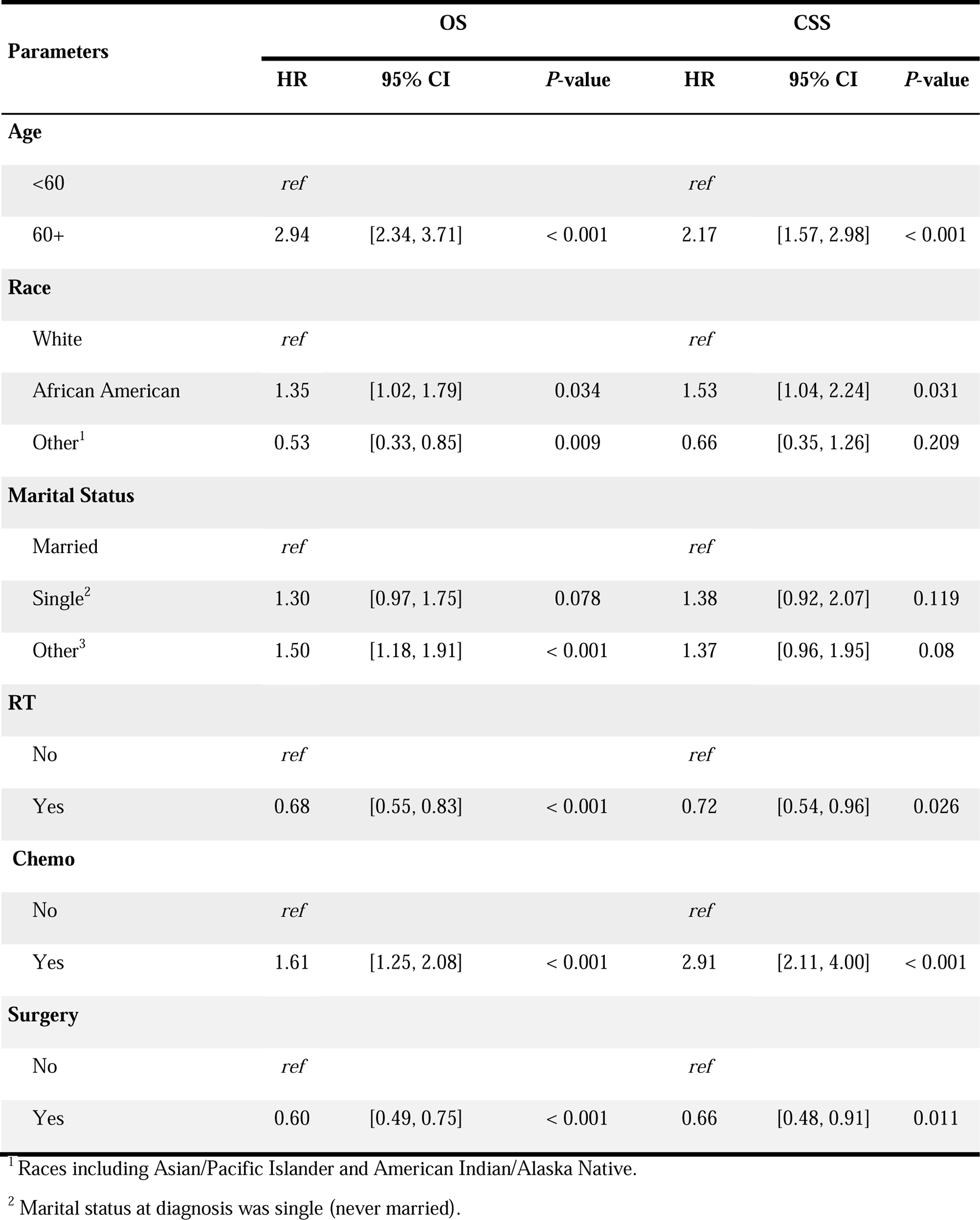
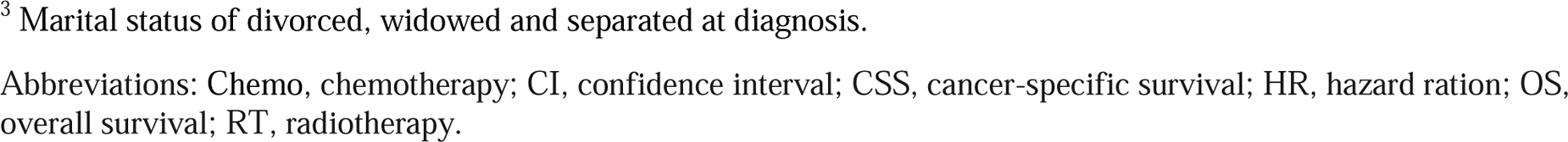
Multivariable cox regression analysis for disease-specific survival of primary extraosseous plasmacytoma patients

| Parameters | OS |  |  | CSS |  |  |
| --- | --- | --- | --- | --- | --- | --- |
|  | HR | 95% CI | P-value | HR | 95% CI | P-value |
| **Age** |  |  |  |  |  |  |
| <60 | *ref* |  |  | *ref* |  |  |
| 60+ | 2.94 | [2.34, 3.71] | < 0.001 | 2.17 | [1.57, 2.98] | < 0.001 |
| **Race** |  |  |  |  |  |  |
| White | *ref* |  |  | *ref* |  |  |
| African American | 1.35 | [1.02, 1.79] | 0.034 | 1.53 | [1.04, 2.24] | 0.031 |
| Other<sup>1</sup> | 0.53 | [0.33, 0.85] | 0.009 | 0.66 | [0.35, 1.26] | 0.209 |
| **Marital Status** |  |  |  |  |  |  |
| Married | *ref* |  |  | *ref* |  |  |
| Single<sup>2</sup> | 1.30 | [0.97, 1.75] | 0.078 | 1.38 | [0.92, 2.07] | 0.119 |
| Other<sup>3</sup> | 1.50 | [1.18, 1.91] | < 0.001 | 1.37 | [0.96, 1.95] | 0.08 |
| **RT** |  |  |  |  |  |  |
| No | *ref* |  |  | *ref* |  |  |
| Yes | 0.68 | [0.55, 0.83] | < 0.001 | 0.72 | [0.54, 0.96] | 0.026 |
| **Chemo** |  |  |  |  |  |  |
| No | *ref* |  |  | *ref* |  |  |
| Yes | 1.61 | [1.25, 2.08] | < 0.001 | 2.91 | [2.11, 4.00] | < 0.001 |
| **Surgery** |  |  |  |  |  |  |
| No | *ref* |  |  | *ref* |  |  |
| Yes | 0.60 | [0.49, 0.75] | < 0.001 | 0.66 | [0.48, 0.91] | 0.011 |

<sup>1</sup> Races including Asian/Pacific Islander and American Indian/Alaska Native.
<sup>2</sup> Marital status at diagnosis was single (never married).
<sup>3</sup> Marital status of divorced, widowed and separated at diagnosis.
Abbreviations: Chemo, chemotherapy; CI, confidence interval; CSS, cancer-specific survival; HR, hazard ration; OS, overall survival; RT, radiotherapy.

### Establishment and Evaluation of Nomogram Models

By integrating age, race, marital status at diagnosis, radiotherapy, chemotherapy, and surgery, a nomogram was constructed to predict the 3- and 5-year OS probability of PEP patients (Figure 5A). The calibration curve of the nomogram revealed a close match between the predicted and observed OS probability at the 3- and 5-year intervals (Figure 5B). Additionally, time-dependent ROC curve analyses indicated that the 3-year and 5-year AUC of the nomogram was 0.746 and 0.738, respectively (Figure 5C), indicating the high accuracy of the nomogram models. The DCA curves also suggested that the nomograms had good performances as effective tools for predicting OS probability (Figure 5D-E). Furthermore, similar results were obtained for the nomograms designed to predict the 3- and 5-year CSS probability of primary extraosseous plasmacytoma patients (Supplementary Figure S1, A-E).

**Figure 5.**
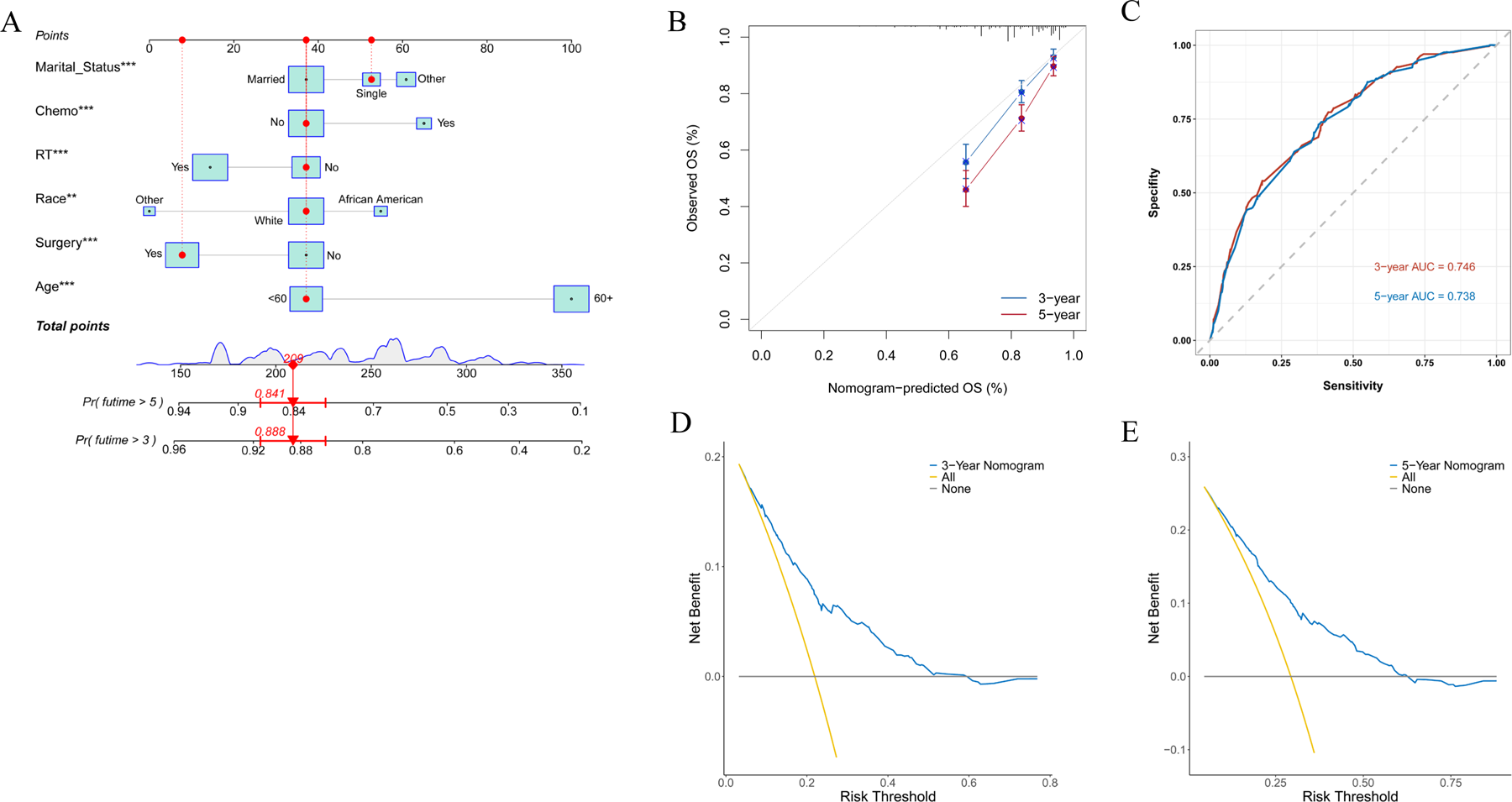
Construction and evaluation of the nomogram of primary extraosseous plasmacytoma patients. (A) The nomogram for predicting the probability of 3- and 5-year OS. The total points were calculated by integrating scores related to age, race, marital status, chemotherapy, radiotherapy, surgery and projected to the bottom scale to predict the OS survival probability at 3 and 5 years. (B) The calibration curve of the nomogram for the observed overall survival (OS) probability and predicted OS at 3-year and 5-year. (C) Time-dependent ROC curve analyses of the nomograms for the 3 years and 5 years in the primary extraosseous plasmacytoma cohort. (D-E) The decision curve analyses of the nomogram for the 3 years (D) and 5 years (E) in the primary extraosseous plasmacytoma cohort. \*\**P* < 0.01, \*\*\**P* < 0.001.

## Discussion

Primary extraosseous plasmacytoma (PEP) is a rare type of plasma cell neoplasm that originates outside of the bone marrow and infrequently converts to multiple myeloma^17^. Due to its rarity, there is limited published literature on PEP, mostly comprising case reports or small case series^18, 19^, which still presents a diagnostic and therapeutic challenge and highlights the lack of experience clinicians have with this condition. The present study identified 1044 PEP patients from 2000-2019 using the SEER database, representing the largest cohort describing the clinical characteristics and outcome of patients with PEP so far. The average age of the patients was 60.3 ± 15.2 years, with a male predominance (male: female = 1.8:1), and the upper aerodigestive tract was the most commonly affected site (42.6%), which is consistent with previous research that has shown extraosseous plasmacytomas to be more prevalent in males of advanced age, typically manifesting as lesions in the upper aerodigestive tract^6, 17, 19^.

Although age has been found to have a significant correlation with the prognosis of multiple myeloma and solitary plasmacytoma in previous studies^20, 21^, its relationship with the survival outcome of PEP has been rarely reported. In the current study, it was found that older age was significantly associated with worse OS and CSS in PEP patients. The 5-year OS and CSS rates were found to decline remarkably with increasing age, especially for patients over 60 years old. Further analysis using both univariate and multivariate Cox regression models confirmed that age was an independent negative prognostic factor for both OS and CSS in PEP patients. This could be due to older patients having more substantial comorbidities, being treated with less aggressive therapies, being less involved in clinical trials compared to younger patients, and being more prone to progression to multiple myeloma due to the effects of aging.

A prior study demonstrated that extraosseous plasmacytoma patients who were African American had the worse survival outcome^22^. Our study yielded similar results, with African Americans having the worst OS, while Asians, American Indians, Native Americans, and Pacific Islanders had the best OS. Further investigation revealed a 1.45-fold hazard ratio in OS and 1.53-fold hazard ratio in CSS for African American patients in relative to White patients after multivariate cox analysis by adjusting for other confounding variables. In contrast, the hazard ratio was 0.45 in OS for the Asians, American Indians, Native Americans, and Pacific Islanders compared to White PEP patients. The disparities may be attributed to the difference in genetic background, dietary culture, and socioeconomic status among those ethnicity groups.

Marital status has been increasingly recognized as an important prognostic factor for cancer patients ^23–25^. Our study indicated that PEP patients with the marital status of “divorced/widowed/separated” had remarkably worse survival outcomes compared to married or single patients. However, there was no significant difference in OS or CSS between married and single patients. After adjusting for other confounding variables through multivariate cox analysis, the hazard ratio for OS was 1.50-fold and for CSS was 1.37-fold higher for “divorced/widowed/separated” patients compared to married ones. The underlying mechanism of shortened survival in “divorced/widowed/separated” patients is unclear, but it may be related to socioeconomic and psychological status, as these patients may have experienced stronger fluctuations in socioeconomic and emotional changes.

Extraosseous plasmacytomas are usually treated with local interventions, such as radiation therapy or surgery, due to their local nature^6^. Surgery may be considered to completely remove extraosseous plasmacytomas in anatomic sites if feasible^14^. However, the upper respiratory tract is most commonly affected site for patients with extraosseous plasmacytoma, it is almost impossible to remove the tumor completely by surgery^26, 27^, in such cases, radiotherapy is often the preferred treatment option^28^. Radiotherapy has been shown to provide excellent local control of solitary plasmacytomas with a lower rate of local relapse^28, 29^. Moreover, surgery can also be employed to avoid long-term sequelae of radiation^14^. There is no clear recommendation for adjuvant chemotherapy for extraosseous plasmacytomas, although it may be considered for larger tumors after radiation therapy^6, 30^. Studies have shown that chemotherapy has no beneficial effect on disease control or prevention of progression to multiple myeloma, and several multicenter studies have shown that chemotherapy does not benefit the survival of patients with solitary plasmacytoma^6, 30, 31^. Our study showed that PEP patients with chemotherapy showed significantly worse survival outcome, while patients with radiotherapy or surgery presented remarkably better survival outcome. Moreover, univariate and multivariate cox analysis showed that radiotherapy and surgery were significantly associated with better prognosis of PEP patients, whereas chemotherapy was related with worse prognosis. This may be related to the fact that patients treated with chemotherapy may be too frail to receive surgery/radiotherapy, or have a more advanced form of disease, requiring more aggressive treatment.

It has become common in clinical practice to use nomograms for predicting cancer prognosis^32^. This study identified age, race, marital status, chemotherapy, radiotherapy, and surgery as independent prognostic factors for the survival outcomes of PEP patients. Nomograms were developed based on these variables to predict 3- and 5-year survival. The nomogram’s calibration curve showed good consistency between predicted and actual survival probabilities, and the nomogram models demonstrated high accuracy, as indicated by their significantly higher AUC. Additionally, the DCA curves indicated that the nomograms were effective tools for predicting the OS probability.

However, there are some limitations with this study. Firstly, the SEER registry did not document other potential prognostic factors that may have a significant impact on PEP patient outcomes, such as LDH level, immunophenotypic features, M protein in blood/urine, carcinogen exposure, family history, alcohol/smoking consumption history, and Epstein-Barr virus status. Secondly, detailed information about therapy was not recorded in the SEER database, making it impossible to analyze the effect of different treatment regimens. Thirdly, this is a retrospective study, which means that there may be unavoidable potential biases. Finally, while the nomograms of primary PEP were constructed and verified using the same database, they were not further validated using another independent dataset. Thus, although this study provided important insights on PEP due to the rarity and lack of large-scale trials of the disease, the results should still be interpreted with caution.

## Conclusions

In conclusion, PEP is a rare disease and the outcome for patients with PEP depends on several factors including age, race, marital status, and treatment options such as chemotherapy, radiotherapy, and surgery. Age, race, marital status, chemotherapy, radiotherapy, and surgery were all identified as independent predictors of OS or CSS for PEP patients. Patients who were younger, Asian or Pacific Islander, American Indian or Native American, single or married, and received radiotherapy or surgery had a more favorable prognosis. Conversely, patients who underwent chemotherapy had a poorer prognosis. To our knowledge, this study represents the largest population-based cohort investigating the clinical characteristics and survival outcome of PEP patients, which can also provide valuable insights into the management of PEP.

## Data Availability

The data analyzed in this study are from the SEER database (https://seer.cancer.gov/) that are available to the public.

## Author Contributions

F.W. came up with the conception, design, data analysis and manuscript preparation.

## Acknowledgments

The interpretation of the data is the sole responsibility of the author(s). The author(s) acknowledge the efforts of the National Cancer Institute and the Surveillance, Epidemiology, and End Results (SEER) Program tumor registries in the creation of the SEER database.

## Abbreviations

EOP: extraosseous plasmacytoma
Chemo: chemotherapy
CI: confidence interval
COD: cause of death
CSS: cancer-specific survival
HR: hazard ration
OS: overall survival
RT: radiation therapy
SEER: Surveillance, Epidemiology, and End Results
SPMs: second primary malignancies
NOS: not otherwise specified
PEP: primary extraosseous plasmacytoma.

## Supplementary Figure Legends

**Figure S1. Construction and evaluation of the nomogram of primary extraosseous plasmacytoma patients.** (A) The nomogram for predicting the 3- and 5-year CSS probability. The CSS survival rates at 3 and 5 years can be predicted by integrating scores related to age, race, chemotherapy, radiotherapy, surgery and projected to the bottom scale to predict the OS survival probability at 3 and 5 years. (B) The calibration diagram of the nomogram for the observed CSS probability and predicted CSS at 3-year and 5-year. (C) Time-dependent ROC curve analyses of the nomograms in the 3 years and 5 years in the primary extraosseous plasmacytoma cohort. (D-E) The decision curve analyses of the nomogram for the 3 years (D) and 5 years (E) in the primary extraosseous plasmacytoma cohort. CSS, cancer-specific survival. \*\**P* < 0.01, \*\*\**P* < 0.001.

