## Supplementary figures and images for "Epidemiology and Survival of Primary Extraosseous Plasmacytoma: Insights from A Population-Based Study with A 20-Year Follow-Up"

### Supplementary Figures

#### Slide 1
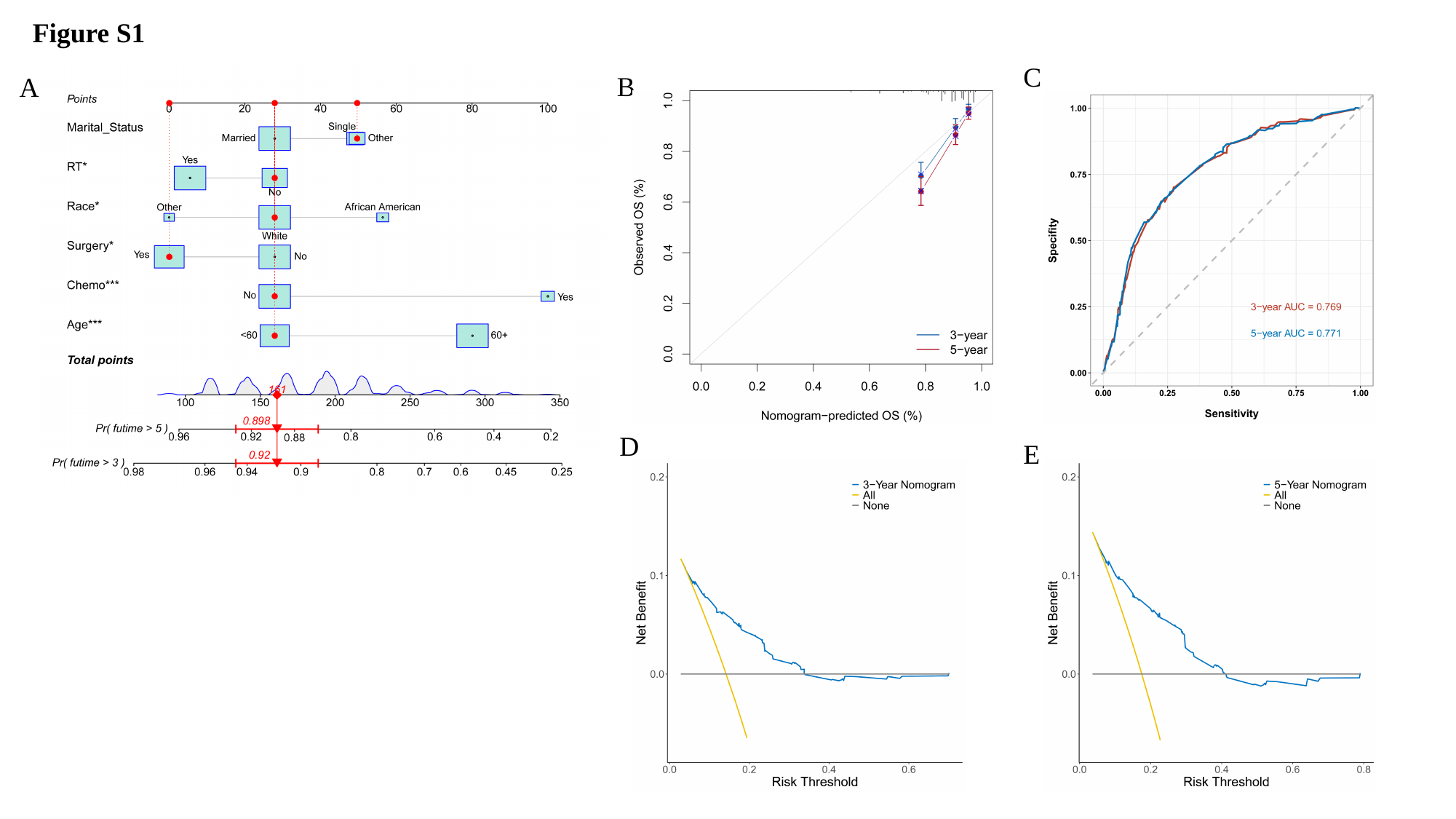

Figure S1
C
B
A
D
E
